# INTEGRATED NCDs TREATMENT AND CARE AMONG PEOPLE LIVING WITH HIV/AIDS ATTENDING CARE AND TREATMENT AT A HEALTH FACILITY

**DOI:** 10.1101/2023.02.21.23286233

**Authors:** Atuganile Musyani, Grace Mosi, Erik Kinyenje, Albertino Damasceno, Mucho Mizinduko, Rogath Kishimba, Meshack Shimwela, Leonard Subi

## Abstract

**Introduction:** Persons living with HIV (PLHIV) in Tanzania now live longer due to the advancement of HIV care programs. For this reason, they are at an increased risk of developing Non-Communicable Diseases (NCDs). Despite many resources committed to HIV care programs, NCDs care is not effectively integrated into these programs. The study aimed at describing missed opportunities to diagnose and manage hypertension and diabetes and implementing and evaluating the effect of three months of preventive efforts among among PLHIV attending care and treatment centers (CTC)

**Materials and methods:** We evaluated 333 PLHIV attending CTC for blood pressure and blood sugar levels. Patients who were diagnosed with high blood pressure equal to or above 140/90 mm Hg, or on treatment for hypertension and a fasting blood sugar above 7.0 mmol/L (126 mg/dl) were subjected to a small intervention aimed at increasing health literacy on the adherence to and control of their pathologies. Re-evaluation of their blood pressure and blood sugar levels was done at the end of a 3 months intervention.

**Results:** Of the evaluated 333 PLHIV, 71 (21.32%) had hypertension and 9 (2.70%) had high blood sugar. Among 177 PLHIV who never had their BP measured before, 37/177 ((20.90%) were diagnosed with hypertension. The cohort group involved analysis of 76 patients with either uncontrolled hypertension or diabetes followed for three months. By the end of the 3^rd^ month of intervention, 26/71 (36.6%) were able to control their blood pressure (BP < 140 SBP and < 90 DBP). The mean blood pressure decreased from 164/99.5 mmHg to 159 /96mmHg. Of the 9 PLHIV with high blood glucose levels, 5/9 (55.5%) had their blood sugar normalized at the end of the intervention. In a focused group discussion, most patients reported difficulties in controlling their BP due to the high cost of medication and consequently poor adherence to medication.

**Conclusion:** The burden of undiagnosed NCDs among PLHIV attending care and treatment clinic is remarkably high. Interventions to address such diseases in CTC seem to work. HIV care and treatment programs should provide integrated care that includes NCDs care.

## INTRODUCTION

Global Non-Communicable Diseases (NCDs) burden remains unacceptably high (1). In 2016, NCDs were responsible for 41 million of the world’s 57 million deaths (71%), where 44% were due to cardiovascular diseases (CVD) and 4% were due to diabetes (2). CVD and diabetes caused 31% and 3% respectively of all deaths globally (3). Deaths due to NCDs are expected to increase to 55 million by 2030 (4). In Tanzania in 2016, among 409,000 deaths, 134,970 (33%) were due to NCDs (5). Of these, (17,546) 13% and (2700) 2% were due to CVD and diabetes mellitus, respectively (6).

Due to the advancement of HIV care programs, persons living with HIV (PLHIV) now live longer (7). For this reason, they are at increased risk of developing Non-Communicable Diseases (NCDs) such as hypertension (HTN) and type 2 diabetes mellitus (T2DM) like other persons due to aging. In addition to the increased risk due to aging, the HIV disease itself and anti-retroviral treatment (ART) use add more risk of NCDs to PLHIV (8–10). Chronic immune activation due to the virus and medication side effects are critical mechanisms that lead to this increased risk (11). Cardiovascular disease is the leading cause of non-AIDS-related morbidity and mortality among PLHIV (11). Diabetes affects PLHIV almost twice as often as persons without HIV (12–14). Patients with multiple comorbidities can benefit by providing integrated care for all the diseases at a single point of care (15–17). Different studies have shown that an integrated platform of care is very crucial in addressing NCDs (17–20).

The prevalence of HIV in Tanzania in 2018 was estimated to be at 4.6% (21). This translated into about 1.4 million children and adults living with the infection in the country. There has been a progressive decline in the number of AIDS-related deaths in the past decade with overall AIDS-related mortality decreasing by 50%, from 48,000 deaths in the year 2010 to 24,000 deaths in the year 2018 (21).

HIV care programs have many resources in contrast with NCDs care that is poorly financed and is not effectively integrated into the HIV care programs (22,23). The country experiences a lack of evidence-based care model for scaling up integrated HIV/NCDs care (2). This is due to inadequate or absence of effective monitoring and evaluation systems (poor NCD surveillance) and inadequate use of health information regarding NCDs (15). The country has weak chronic disease care health system which is not patient-centered, resulting in ineffective control of chronic diseases among all and also among PLHIV (15). There are often inadequate multi-sectoral responses to the diseases, inadequate resources (human, infrastructures, and funds), poor governance and leadership, and low capacity of health service providers in terms of knowledge, skills, and numbers (22).

Our project, therefore, aimed at describing missed opportunities to diagnose and care for hypertension (HTN) and diabetes (DM) among PLHIV attending CTC in Dar es Salaam, Tanzania, and implementing and evaluating hypertension and diabetes mellitus preventive efforts among these patients.

## MATERIALS AND METHODS

### Project design

This was a facility-based screening program for hypertension and diabetes among PLHIV receiving care at CTC in Dar es Salaam, Tanzania. It was conducted from January to June 2020.

### Project setting

The project was carried out at CTC at Temeke Regional Referral Hospital (Temeke RRH). Temeke RRH is a public owned hospital that is located in the southern part of Dar es Salaam region, the largest commercial city located in the Eastern part of Tanzania. Although the hospital has specialized diabetic, cardiovascular, and CTC clinics, the services are not integrated. Patients who have been diagnosed with HTN or diabetes in an HIV clinic are referred for care in these separate clinics within the hospital settings.

### Target population

The target population was PLHIV aged 18 years or above attending Temeke RRH in Temeke District in Dar es Salaam, Tanzania. We used convinient sampling to select PLHIV who attended CTC within the 5 days of data collection. Clients were randomly selected based on availability at the time of interview. The project excluded pregnant women living with HIV/AIDS and patients with mental diseases.

### Project variable

Data collected involved demographic information, blood pressure and biochemical measurements (Blood glucose level and cholesterol level). The project also involved a short discussion for exploring the factors leading to poor adherence to hypertensive and diabetic medication. The primary outcome at various time points were missed opportunities for diagnosis and care for NCDs among PLHIV. Secondary outcomes were control rates of hypertensive and diabetic patients at the end of a three month intervention.

### Blood pressure and glucose measurements

Blood pressure and glucose level were measured following the Tanzania national guidelines. Blood pressure (BP) was measured by trained personnel using a standardized protocol and validated and regularly calibrated electronic sphygmomanometers (Omron HEM-7270). Blood sugar levels were measured using rapid methods (Accu-chek®). Fasting and random blood sugar level was used and diagnosis was based on Tanzania national guideline. Fasting blood glucose was considered when patient reported not eating for more than 8 hours. Blood pressure was checked three times, 5 minutes after arrival to the clinic, during triaging at the triage desk (approximately 30 minutes later), and at the consultation desk with a doctor. The mean of the three measurements of blood pressure were used for diagnosis of hypertension. Hypertension and T2DM were defined according to the Tanzania national guideline (9). Hypertension was defined as blood pressure (BP) equal to or above 140/90 mmHg, T2DM was defined as raised fasting blood sugar of 7.0 mmol/L (126 mg/dl) or more or a random blood sugar equal or above 11.1 mmol/L (200 mg/dL). Blood pressure control was defined as when the systolic BP was below 140mmHg and diastolic BP below 90 mm Hg on anti hypertension treatment. DM was defined as being controlled when fasting blood glucose was below 6 mmol/L.

### Enrolled in a follow-up group

A cohort of 76 patients with HTN and T2DM/high blood glucose levels during the current project was followed for three monhs. At each clinic visit, patients received health education on hypertension and DM. Individuals with hypertension were managed using a clinical and medication management algorithm based on the Tanzania clinical guideline (9). Participants diagnosed with diabetes were encouraged to do a lifestyle change. The patient’s registry was developed following recommendations from the HEARTS technical package of the World Health Organization (WHO) and Tanzania national guideline with some modification (10,11). A simple surveillance system was developed for all patients diagnosed with HTN and T2DM. All 76 patients had complete data and were involved in the analysis at the end of the study.

**Figure 1.** The diagram showing the PLHIV diagnosed with hypertension, DM or both and enrollment into the cohort group for follow-up.

### A focus group discussion

A focus group discussion was conducted to ascertain participant knowledge about BP control and reasons for poor blood pressure control. A convenience sample of patients was identified that included 10 clients and 10 health care workers.

### Analysis of data

Data were entered and cleaned using Epi info version 7, and then analyzed using STATA version 15 for Windows (Stata Corp., College Station, TX, USA). Descriptive statistics such as median (with interquartile range) and frequencies (with percentages) were used to illustrate the baseline characteristics of participants with the relevant variables. A short description of qualitative information was reported. Missing data were excluded from the analysis. Chi-square test was used to compare categorical variables between intervention and non-intervention group in relation to hypertension and diabetes diagnosis outcome. Fisher’s exact test was used where appropriate. The null hypothesis was that, there was no difference in distribution of characteristics between intervention and non-intervention group. An alternative hypothesis was that, at least one characteristics was related to intervention and non-intervention group. Binary logistic regression was used to ascertain association between the independent and dependent variables. All independent variables with a P value ≤ 0.1 in the bivariate analyses were included in the final multivariable logistic model in order to identify the risk factors for getting hypertension and diabetes. All variables which attained a p-value <0.05 was considered statistically significant. A paired t-test was conducted on the measurements of 76 participants (systolic and diastolic pressure, BMI, and fasting blood glucose levels) to determine if measurements at month three were different from those taken at baseline to ascertain the effectiveness of the intervention.

### Ethical consideration

An ethical clearance to conduct this project was obtained from the Muhimbili University of Health and Allied Sciences Institutional Research Board. Permission to conduct the study at Temeke Regional Referral Hospital was obtained from the Medical Officer In-charge. All participants gave informed consent to participate in the project after a thorough description of the study objectives and risks associated with participation. The project ensured privacy and confidentiality to all participants.

## RESULTS

### 3.1. Demographic and Clinical Characteristics of the Study Participants

#### Study size

A total of 335 PLHIV participated in the interview, with two (2) participants removed from the analysis due to incomplete information. A total of 333 patients were included in the evaluation for hypertension and blood sugar levels. Demographic and clinical characteristics were compared between patients diagnosed with diabetes and hypertension (intervention group) and those not diagnosed with either diseases (control group) as shown in *table 1*. The median age (IQR) for the study participants was 46 (39-52) years. There was a significant age difference between intervention and control group (*P-value 0*.*02*). The intervention group were a bit older with a median age (IQR) of 48 (42-58) compared to the control group median (IQR) age of 46 (38-51). Except for age, body mass index and waist hip ratio which showed a significant difference between intervention and control group, there no significant difference in all other variables between the two groups (*Table 1*).

**Table 1:**
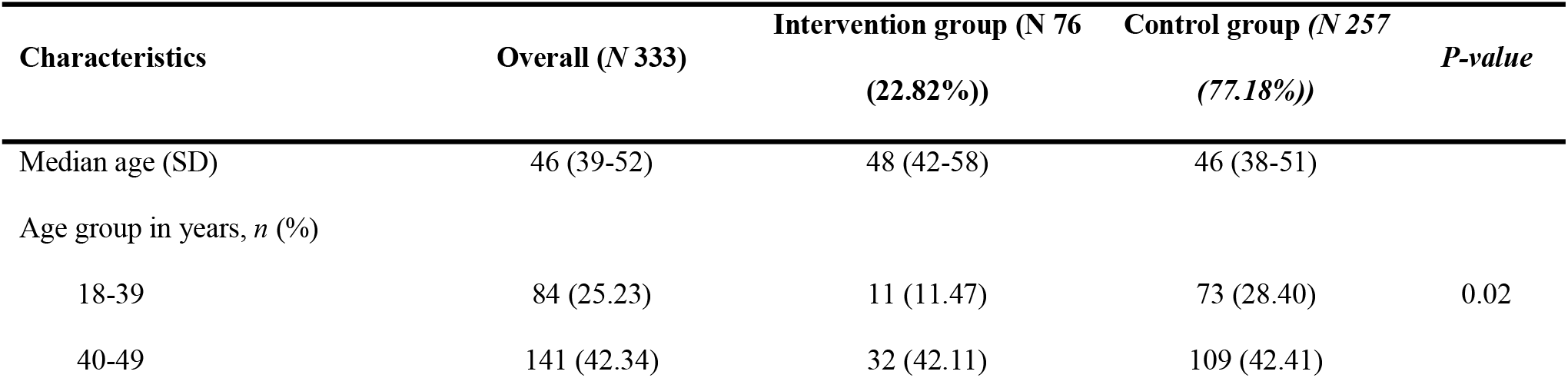
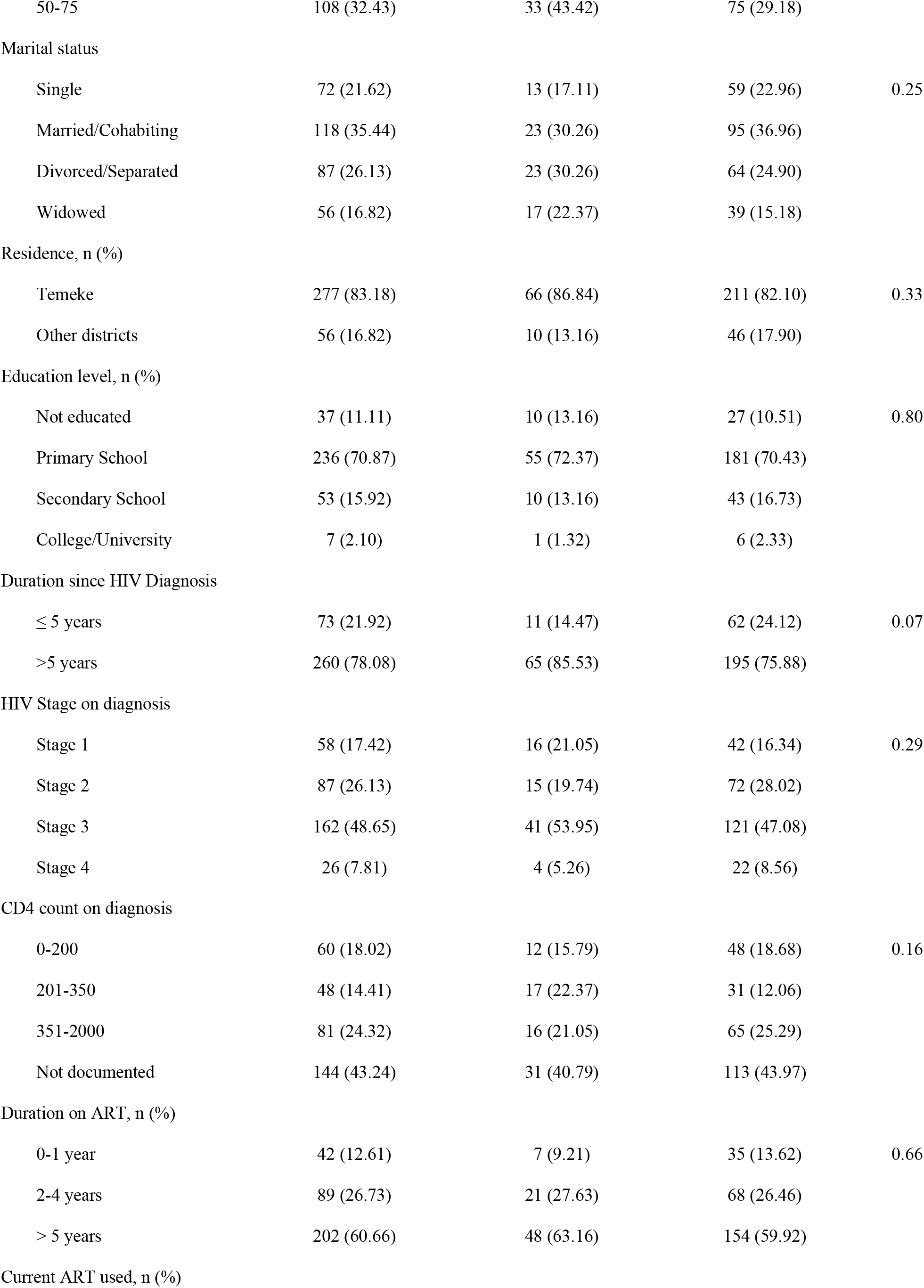
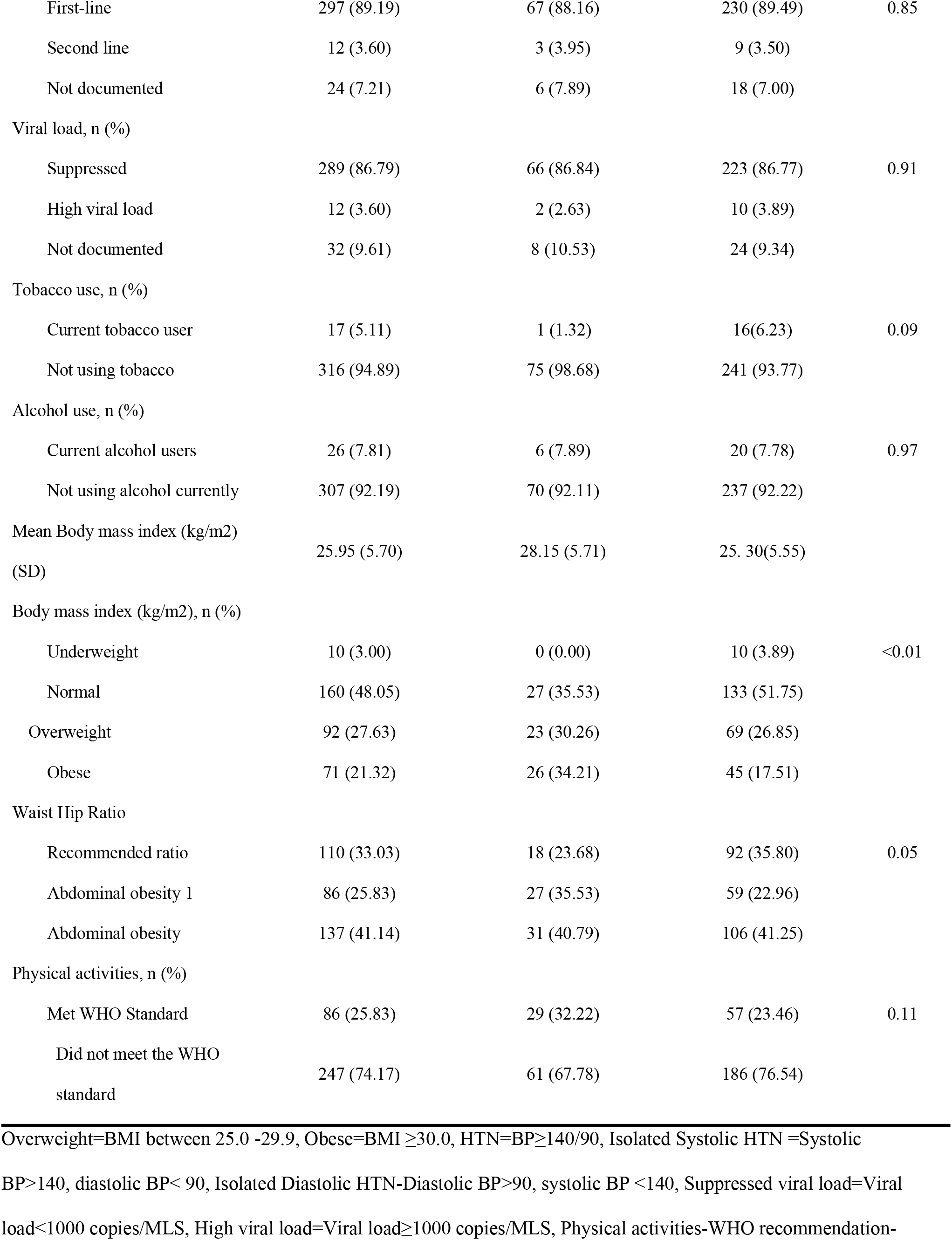
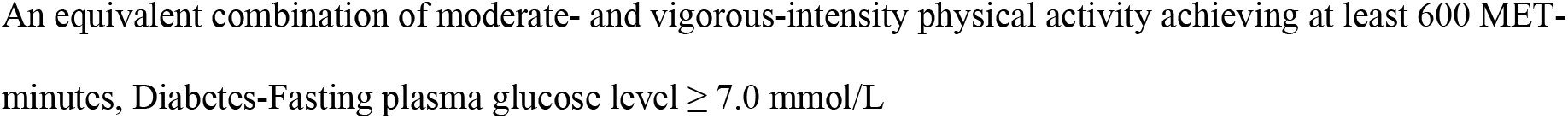
Demographic and Clinical characteristics of patients diagnosed and not diagnosed with hypertension and diabetes among people living with HIV/AIDS attending Care and Treatment Center in Dar es Salaam, Tanzania in 2020.

Overall, of 333 participants, 71/333 (21.32%) participants had hypertension and 9/333 (2.70%) were diabetic. Four 4/333 (1.20%) participants were diagnosed with both hypertension and diabetes. Of the 71 patients with hypertension, only 34/71 (47.89%) knew they had hypertension and only 5 (7%) were already on treatment but had not achieved blood pressure control. Over half of the study participants 177/333 (53.15%) have reported never having their blood pressure measured before, and among these 37/177 (20.90%) were newly diagnosed with hypertension. Overall, of the 9 participants with diabetics, 4/9 (44.44%) knew they had diabetes and were on treatment and 5/9 (66.67%) were diagnosed during the study.

The prevalence of hypertension was significantly higher in patients older than 40 years (32/71 (42.11) and 50 years (33/71, 43.42%) compared to the younger counterparts aged less than 39 years (11/71, 11.47%; P<0.05).

### Univariate and Multivariate analysis

Univariate and multivariate analyses results are shown in Table 2. All variable with crude OR of equal or more that 0.1 in the univariate analysis were subjected to multivariate analysis. In the analysis of risk factors, obese patients were 4 times as likely to have hypertension (AOR = 3.67; 95%CI: 1.64 – 8.17) compared to patients with normal body weight. This finding was significant even after controlling for other confounder including age, gender. People above 40 and 50 years were twice at risk each of getting hypertension compared to people aged below 40 years (AOR 1.76 95%CI 0.71-3.35 and AOR 2.48 95%CI 0.96-6.42 respectively). People with diabetes mellitus were 5 times at significant risk of getting hypertension although the sample size was not adequate for comparison (AOR 5.30 95%CI 1.04-26.98).

**Table 2:**
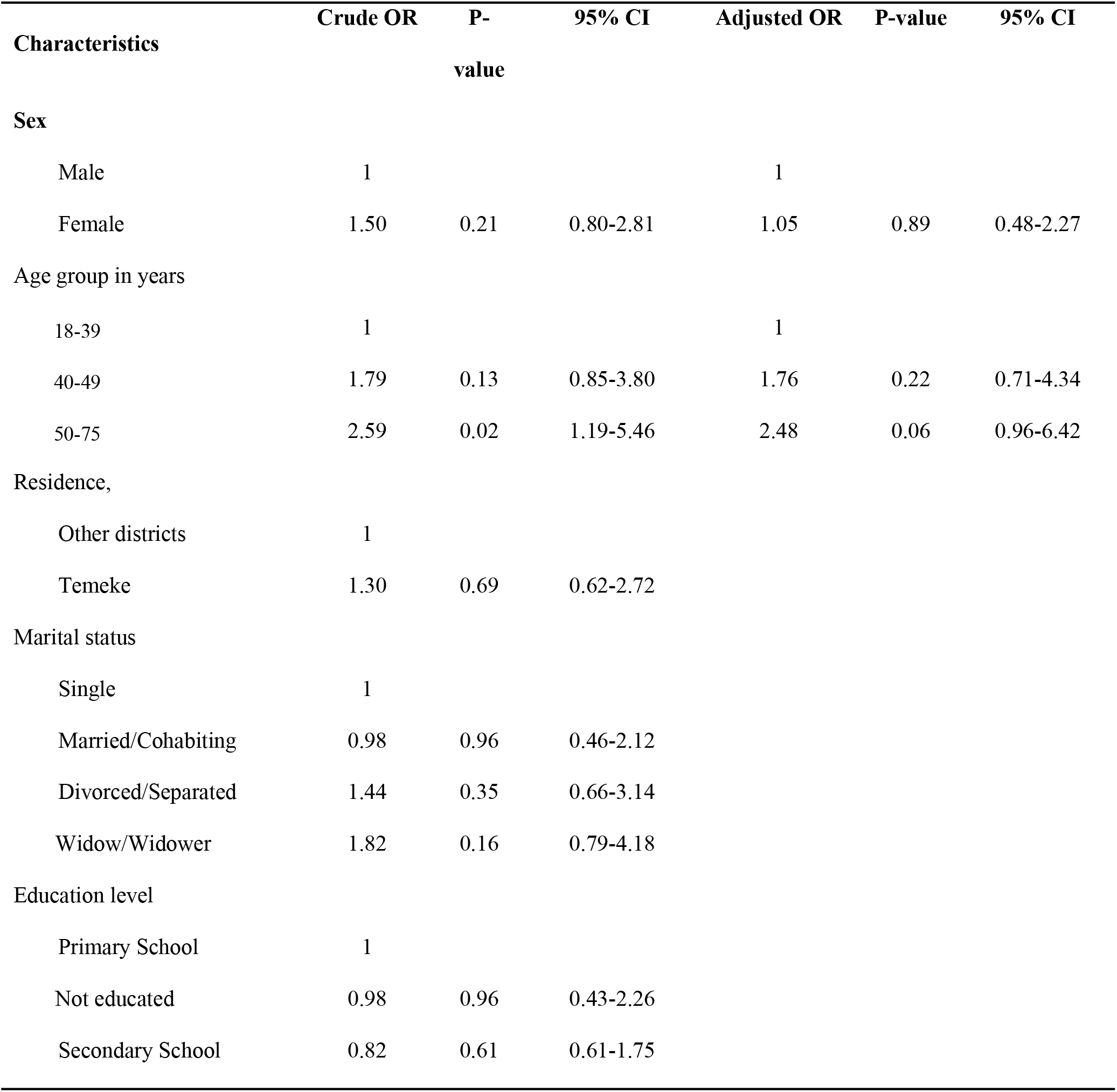
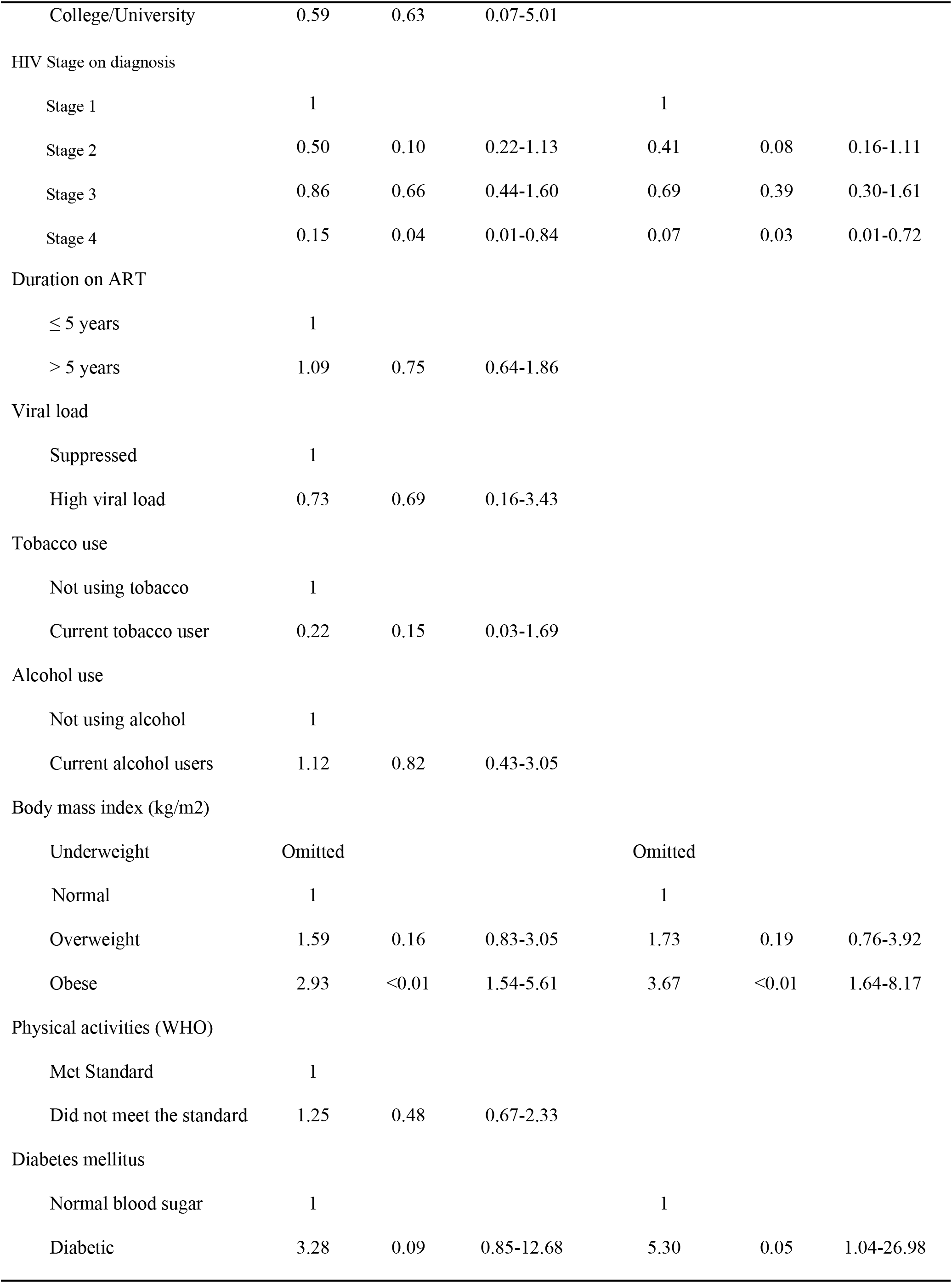
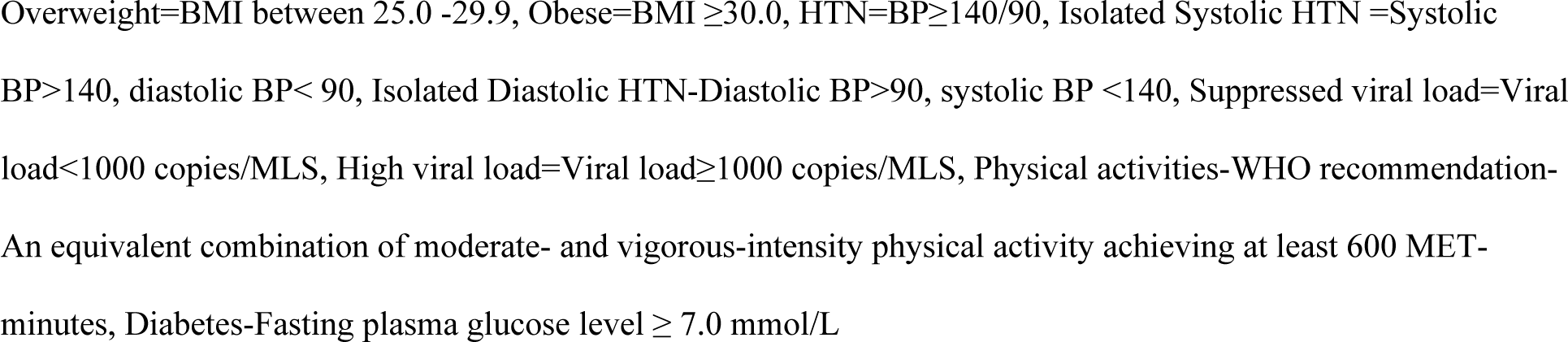
Association between risk factors and elevated blood pressure or blood glucose among people living with HIV/AIDS attending Care and Treatment Center in Dar es Salaam, Tanzania in 2020.

All patients included in the cohort (76) had completed data and were available for analysis at the end of the 3 month study.

After three months of intervention, the mean change in blood pressure for study participants was from 164 to 159 mm/Hg for systolic blood pressure and 100 to 96 mm/Hg for diastolic blood pressure. Blood sugar level of patients who were diagnosed to be diabetic changed from the mean value of 11.96 to 9.38 mmol/L. Also for BMI, the change was from 28.9 to 27.5 kg/m2. ***(Table 3)***

**Table 3:**
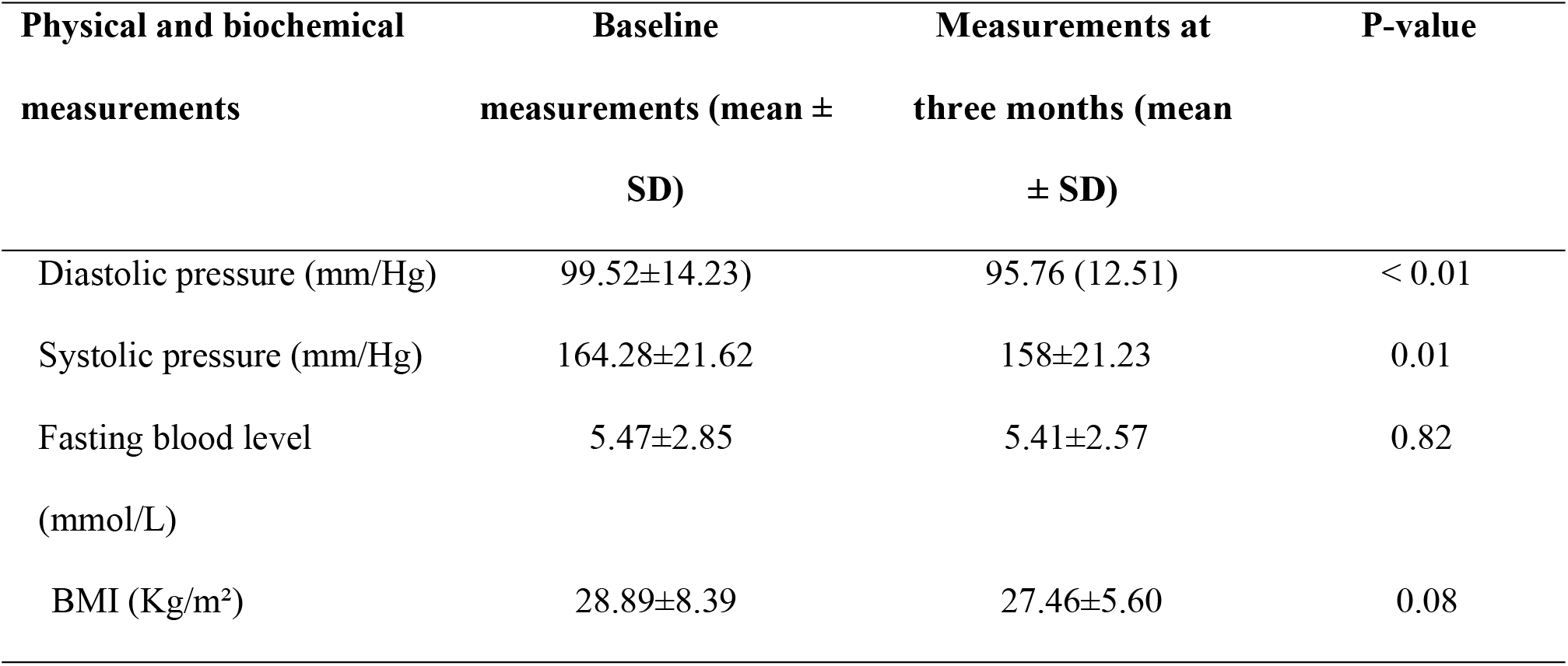
The effects of three months of preventive efforts in 76 HIV positive patients with elevated blood pressure or blood glucose attending Care and Treatment Center in Dar es Salaam, Tanzania in 2020.

| Physical and biochemical measurements | Baseline measurements (mean ± SD) | Measurements at three months (mean ± SD) | P-value |
| --- | --- | --- | --- |
| Diastolic pressure (mm/Hg) | 99.52±14.23) | 95.76 (12.51) | < 0.01 |
| Systolic pressure (mm/Hg) | 164.28±21.62 | 158±21.23 | 0.01 |
| Fasting blood level (mmol/L) | 5.47±2.85 | 5.41±2.57 | 0.82 |
| BMI (Kg/m<sup>2</sup>) | 28.89±8.39 | 27.46±5.60 | 0.08 |

Results for a paired t-test showed a significant decrease in mean systolic blood pressure (t = 3.07 w/df=81, p <0.01, 95%CI (2.01, 9.45)) and mean diastolic blood pressure (t=2.91 w/df=81, p<0.01, 95%CI (1.19, 6.33)). There was no statisticaly significant difference in mean FBG level (t=0.03 w/df=81, p =0.82, 95%CI (−0.45, 0.57)) and BMI (t=1.76 w/df=81, p =0.08, 95%CI (−0.18, 3.03)).

Univariate and multivariate analysis were conducted to show the relationship of factors at the start of intervention with the decrease in blood pressure and blood glucose level. The factors to include in the univariate and multivariate analysis were selected based on being significant risk factors for hypertensive or diabetic. There were no factors related to decrease in blood pressure and glucose blood level at the univariate analysis, although at multivariate analysis, age above 50 years and being obese at the start of intervention were related to decrease in blood pressure level *Table 4*.

**Table 4:** Association between risk factors and decreased blood pressure or blood glucose among 76 people living with HIV/AIDS after three months of intervention in a Care and Treatment Center in Dar es Salaam, Tanzania in 2020.

| Characteristics | Crude OR | P-value | 95% CI | Adjusted OR | P-value | 95% CI |
| --- | --- | --- | --- | --- | --- | --- |
| **Sex** |  |  |  |  |  |  |
| Male | 1 |  |  | 1 |  |  |
| Female | 1.50 | 0.21 | 0.80-2.81 | 1.22 | 0.55 | 0.63-2.37 |
| **Age group in years** |  |  |  |  |  |  |
| 18-39 | 1 |  |  | 1 |  |  |
| 40-49 | 1.57 | 0.64 | 0.24-10.49 | 1.78 | 0.14 | 0.83-3.82 |
| 50-75 | 0.96 | 0.97 | 0.16-5.93 | 2.27 | 0.04 | 1.04-4.96 |
| **Residence,** |  |  |  |  |  |  |
| Other districts | 1 |  |  |  |  |  |
| Temeke | 0.54 | 1.75 | 0.84—18.0 |  |  |  |
| Marital status |  |  |  |  |  |  |
| Underweight | 1 |  |  | 1 |  |  |
| Normal | 0.66 | 0.49 | 0.20-2.15 | 1 |  |  |
| Overweight | 0.75 | 0.65 | 0.22-2.60 | 1.49 | 0.24 | 0.77-2.88 |
| Obese | Omitted |  |  | 2.56 | 0.01 | 1.30-5.05 |
| Marital status |  |  |  |  |  |  |
| Single | 1 |  |  |  |  |  |
| Married/Cohabiting | 0.98 | 0.96 | 0.46-2.12 |  |  |  |
| Divorced/Separated | 1.44 | 0.35 | 0.66-3.14 |  |  |  |
| Widow/Widower | 1.82 | 0.16 | 0.79-4.18 |  |  |  |
| Physical activities (WHO) |  |  |  |  |  |  |
| Met Standard | 1 |  |  |  |  |  |
| Did not meet the standard | 0.29 | 0.13 | 0.06-1.42 | 1.23 | 0.53 | 0.65-2.32 |

### Adherence to diabetic and hypertensive medication

Three focused group discussions were conducted, 10 participants and 10 healthcare workers were interviewed to explore the understanding of the participants on various risk factors for hypertension and diabetes. Also, barriers to care and adherence were discussed. Pre-defined questions were created to guide the discussion. After enough information was collected the discussion was stopped. Responses were documented during the focus group by one of the project implementers in a paper for reference during report writing.

Almost all participants had good knowledge of hypertension and diabetes. From the discussion, the most reported challenges to adherence to medication were lack of money to buy medication, unaffordable consultation fees, and medication side effects. Most of the participants didn’t know the type of medication regimen they are taking, while others never took medication after diagnosis or even after hypertensive and diabetic disease medication have been prescribed. Few reported taking medication when they develop some hypertensive or diabetic symptoms as most other f time were asymptomatic. The main reason was the high cost of medication or feeling well even after diagnosis. Additionally, other participants were diagnosed and kept on treatment for either HTN or DM other than the current clinic but never reported that they were on treatment. Health care workers reported the absence of a specialized NCDs clinic at the Care and Treatment Center (CTC). They also have no current knowledge of the management of hypertension and diabetes to PLHIV. They have reported having no access to routine physical and biochemichal checkup at the clinic hence rendering them to miss patients suffering from NCDs.

## DISCUSSION

In this project, we found that integration of hypertension (HTN) and diabetes (DM) prevention efforts among PLHIV attending care is suboptimal in a care and treatment clinic. Most PLHIV reported to never measured their blood pressure and some had a silent disease that was not diagnosed at the clinic due to the absence of routine screening of NCDs among PLHIV. The risk of having hypertension among PLHIV was high in obese patients and diabetic patients. The project also revealed that PLHIV with other NCDs co-morbidities do not adhere to NCDs medication due to high medication costs and having a silent disease. Health care workers providing HIV care do not have up-to-date knowledge on hypertension and diabetic care rendering it difficult providing of NCDs care among PLHIV. Close follow-up on hypertension and diabetes among PLHIV was found to be productive due to a significant decrease in mean systolic and diastolic blood pressures at the end of the follow-up period.

In our study, many HIV-positive were not screened for hypertension and diabetes in the facility revealing missed opportunities to diagnose other co-morbidities among PLHIV. Due to the absence of routine screening and care of hypertension and diabetes at the clinic, patients although known to have hypertension did not report on their conditions to health facilities. This is similar to the results found in Malawi and other parts of Africa where there was low integration of NCDs care in PLHIV (15,19,24). This result contradicts results obtained in another study in Tanzania which reveals the integration of care to be practiced in some health facilities with high acceptability of the model (25). The main reason for these findings is probably due to HIV care settings that do not mandate the clinician to provide NCDs care among PLHIV. As this has not been the practice, health care providers in CTC did not have up-to-date information regarding the provision of NCDs care among PLHIV similar to the findings from a study done in South Africa (26). Findings from our study call for the country to ensure health care professionals providing HIV care to be well trained on providing comprehensive care that also includes NCDs care apart from focusing on only HIV services.

Hypertension is a major risk factor for heart diseases however this is not well known among PLHIV (15,27–29). Our project found the prevalence of HTN to be 21.32%. This prevalence is lower compared to the estimated prevalence of 26% in the general population in Tanzania (30). Similar findings were obtained in a study conducted in Senegal (31). The prevalence of hypertension observed in this study is higher compared to a study which was conducted in rural Tanzania which found the prevalence of hypertension to be 11.6% (9,32). However, it was lower than that reported by studies conducted elsewhere in Mbeya and Ethiopia (32,33). The results in this project could be explained due to the nature of the project which relied on convenience sampling.

Obesity was found to be an important risk factor for the development of hypertension in the current study. This finding were obtained in this study suggesting that obesity may lead to the development of hypertension. Similar findings were found in other studies conducted in Tanzania and other countries in Africa (31–33). These finding could be explained by the growing evidence that increased visceral fat in the body is a pathological depot that leadeth to accumulation of more visceral fat in tissues (34,35). Visceral fat have been linked to increased inflammatory reactions in endothelial cells of the blood vessels due to its sensitivity to lipolysis leading to endothelial dysfunction and arterial hypertension (34,35). More evidence from different studies shows that increased prevelance of overweight and obesity has caused increased burden of hypertension (36,37). These finding adds to the evidence that as BMI is easily measured and widely accessible it remains to be a simple and effective tool for screening hypertension in public practice.

In our study, diabetes was also associated with hypertension. Similar findings were found in a study that was conducted in Northeast Ethiopia (33). These findings further add to the evidence that diabetes is a risk factor for hypertension and adds up to the burden of HIV and the need for screening diabetes for all hypertensive HIV-positive patients (38,39).

Furthermore, our project found a significant change in blood pressure after three months of intervention both for systolic and diastolic blood pressure showing the effectiveness of the intervention. These findings were similar to findings in a study conducted in Bangladesh although only physical and behavioral measurements were considered and not biochemical and blood pressure levels (16). Our project showed no significant change in BMI value, while a study in Bangladesh showed a significant change in the level of physical activity which might result in a change in BMI value (16).

This projectwas conducted in an HIV setting where normal clinic visits took place so did not result in disruption of care to PLHIV. Although risk factors were identified and added to the evidence, information cannot be generalized elsewhere in Tanzania but can be used as a model to better research studies and program implementation. Participants were not randomly selected so some findings are prone to bias, this was because the study focused on control program, or prevention program and provide concise documentation to that effect. The other important limitation is that the project was implemented during the COVID-19 pandemic which affected every part of the world. In our study, it has affected the scheduled patient follow-up clinic and measurements as PLHIV were among the group prone to infection hence they were given long clinic visits to avoid congestion at the clinic. Few HCWs were also scheduled in shifts to attend a clinic due to lack of protective gear, hence further hindering consultative services to hypertensive patients. To overcome this we used phone calls to follow-up patients and extension of time for clinic followup to garantie that all 76 clients in the cohort managed to have second test for both blood pressure and blood glucose.

## Data Availability

https://doi.org/10.5061/dryad.05qfttf6x

## CONCLUSION AND RECOMMENDATION

Our project found that most HIV-positive patients are not screened for hypertension or other NCDs. On top of that, we found that health education and close follow-up using surveillance database is of importance for monitoring of patients. We found also that, adherence to antidiabetic and antihypertensive medication is inadequate due to the high cost of medication and lack of follow-up and monitoring. Also, the project found that being obese and diabetic are the risk factors for hypertension. Working in the government of private organization and stage four HIV disease were associated with lower risk for hypertension. Overall, our project has shown that integration of NCDs care in HIV clinics is possible and health education-based intervention at health facilities can result in positive change if practiced effectively. This method can be recommended as a promising strategy in reducing the NCD burden among PLHIV.

We recommend the integration of NCDs and HIV services in Tanzania. Also we recommend the need to strengthen health education and screening of NCDs as the prevalence continues to grow among PLHIV and significant changes are observed with the creation of awareness. The cost of antihypertensive and antidiabetic medication needs to be afordable, especially to PLHIV. We recommend also more robust methodological studies on the integration of NCDs care among PLHIV to make evidence-based information using a big sample size with all inclcusive and exclusive criteria into consideration.

## ACKNOWLWDGEMENT

The authors wish to thank the The Centers for Disease Control and Prevention (CDC) for financial support.

## AUTHOR CONTRIBUTION

Conceptualization: AM, GM, EM. Data curation: AM, GM. Formal analysis: AM, GM, EM. Funding acquisition: AM. Investigation: AM. Methodology:AM, GM, EM. Project administration: AM. Software: AM. Supervision: MM, RK, AT, LB, MS. Validation:MS. Visualization: AM. Writing – original draft: AM. Writing – review & editing: AD,MM,AT

